# Endothelial function is associated with incident diabetes mellitus: the ELSA-Brasil cohort study

**DOI:** 10.1101/2025.04.14.25325841

**Authors:** Karina P. M. P. Martins, Sandhi M. Barreto, Lidyane V. Camelo, Sara T. Menezes, Naomi M. Hamburg, Maria de Fatima H. S. Diniz, Antonio L. P. Ribeiro, Luisa C. C. Brant

## Abstract

**Background:** Diabetes mellitus (diabetes) is associated with the development of endothelial dysfunction. However, the directionality of this association has been debated. Our primary objective was to evaluate the association of microvascular endothelial function endothelial function with incident diabetes in Brazilian adults. We also assessed whether there was an effect modification according to obesity status.

**Methods:** In participants from the ELSA-Brasil cohort study, free of diabetes, and with valid exams for endothelial function at baseline, we evaluated the association between endothelial function and incident diabetes after 8.8 years (SD=2.1) of follow-up. Microvascular endothelial function was assessed by peripheral arterial tonometry (PAT). The variables mean basal pulse amplitude (BPA) representing basal vascular tone, and PAT ratio, the vasodilatory capacity in response to reactive hyperemia, were analyzed in sex-specific quartiles. Higher BPA and lower PAT ratio reflect more impaired endothelial function. Cox proportional hazard models were used for statistical analyses, and multivariable models were adjusted for sex, age, educational level, physical activity level, smoking, alcohol use, and body mass index (BMI) or waist circumference (WC).

**Results:** In 1,268 participants, mean age was 51±8 years, with 45% women, and mean BMI of 26.4±4.4 kg/m^2^. After follow-up, we identified 159 new cases of diabetes. The incidence rates varied from 6.5 (95%CI 4.0-10.2) in the first quartile to 19.7 (95%CI 15.1-25.8) in the fourth quartile for BPA and from 7.2 (CI 95% 4.7-11.1) in the fourth quartile to 15.3 (CI 95% 11.4-20.6) in the first quartile of for PAT ratio. After adjustments, we found that the risk of diabetes increased significantly in those with more impaired endothelial function, for both BPA HR 2.13 (95% CI 1.24 – 3.66, p<0.05) and PAT ratio 1.54 (95% CI 0.90 – 2.60, p 0.11). There was no effect modification according to obesity status, evaluated by BMI and WC.

**Conclusions:** In Brazilian adults, greater impairment of endothelial function at baseline predicted diabetes after 8.8 years of follow-up, independent of obesity status. These findings may have implications for understanding cardiometabolic diseases in the context of pathophysiology, prognosis and treatment.

## INTRODUCTION

Diabetes mellitus is a metabolic disorder associated with an increased risk of all-cause and cardiovascular mortality.^1^ Atherosclerotic cardiovascular disease (CVD) is the major cause of morbimortality in individuals living with diabetes. In 2021, the number of individuals living with diabetes worldwide was approximately 536.6 million, and it is projected to increase to 783.2 million by 2045. Brazil is among the five countries with greater number of individuals with diabetes, with 15.7 million in 2021, and an estimated 23.2 million by 2045, as the prevalence is rising along with the increasing prevalence of obesity.^2–4^ In this context understanding the pathophysiologic mechanisms and the determinants of diabetes are fundamental to advance prevention and reduce the consequences of the disease.

Endothelial dysfunction is an early expression of vascular disease related to cardiovascular risk factors and can predict events.^5^ We and others^6,7,8^ have also previously demonstrated that more impaired microvascular vasodilatory function was cross-sectionally associated with metabolic risk factors, including higher BMI, total cholesterol/HDL and triglycerides. It was also demonstrated that even metabolically healthy individuals with obesity have impaired microvascular function.^9^ Cross-sectional associations between diabetes and impaired endothelial function have been demonstrated by large cohorts, including the ELSA-Brasil,^6,7,8^ but more longitudinal studies are needed to understand the direction of this association.

The ability of endothelial dysfunction to predict incident diabetes has already been demonstrated.^10–14^ This finding is attributed to the effects of hyperglycemia and insulin resistance, which may result in a reduced bioavailability of vascular nitric oxide (NO) and inappropriate production of oxygen-free radicals, culminating in endothelial dysfunction.^15^ However, these studies used diverse methodologies to evaluate endothelial function, some of which are complex and/or invasive, making them difficult to be applied in clinical practice. Only two studies evaluated microvascular function using a non-invasive method in individuals from Europe, where the nutritional transition is in a different stage compared to Brazil.^12,14^ As such, we aim to investigate the association between endothelial function and incident diabetes in the Brazilian context, and evaluate whether there was effect modification of this relation according to obesity status, as a secondary analysis. We hypothesized that endothelial dysfunction would predict diabetes in Brazilian adults, with greater effect size in those individuals living with obesity.

## METHODS

### Study Design and Setting

The study sample derived from the ELSA-Brasil study, a multicenter cohort of 15,105 active or retired civil servants from six universities and research institutes in six Brazilian capitals, aged 35 to 74 years, who volunteered to participate. We collected data during four visits (2008–2010, 2012–2014, 2017–2019, 2022-2024) and through annual telephone surveillance.^16^ Other details of concept, design and cohort profile were published elsewhere. ^16,17^

The current study sample derived from the participants enrolled at the Minas Gerais Investigation Center, where peripheral arterial tonometry (PAT) data acquisition began partway through the baseline visit (2008–2010). Detailed description of patients who performed endothelial function at baseline has been previously published.^8^ From the 1,659 participants who were eligible for PAT exams, 124 participants were excluded due to: Raynaud’s disease (n=1), radical mastectomy (n=3), refusal (n=2), hand abnormality (n=5), acute illness 12 hours before the PAT (n=58), incomplete data (n=2), technically inadequate studies (n=50). Hence, there were 1,535 participants who had valid PAT at baseline. For the current study, all ELSA-MG participants who attended baseline, underwent PAT exam, and were free of diabetes were eligible.

Demographic and clinical variables as age, sex, smoking status, and history of a previous medical diagnosis of diabetes and anti-diabetic medication use were collected through interviews and followed the ELSA-Brasil’s study protocol.^16–18^ In each visit, after an overnight fasting of >8h, we measured weight, height, and waist circumference following internationally standardized protocols. The blood samples were obtained and frozen to be analyzes in a central laboratory. Plasma glucose was measured using the hexokinase method (Cobas c501R, Roche Diagnostics, Rotkreuz, Switzerland) and glycated hemoglobin (HbA1c) by high-pressure chromatography (HPLC—Bio-Rad Laboratories, Hercules, CA, USA).^19^ The study protocol was approved at UFMG’s Institutional Review Board, and all participants signed an informed consent before any study procedure was performed. We followed the STROBE guidelines for cohort studies to report this analysis.

### Explanatory variables: Microvascular endothelial function

Endothelial function was assessed by PAT. PAT exam was performed through Endo-PAT2000 (Itamar Medical Ltd., Caesarea, Israel) by two certified examiners.^8,20^ Briefly, the cuff was placed in the participant’s nondominant arm, 2 cm above the cubital fossa, and PAT probes on each index finger. Baseline pulse amplitude was measured for 5 min. Arterial flow was interrupted on one side for 5 minutes by inflating the cuff at suprasystolic pressure. After 5 min, the cuff was deflated to induce reactive hyperemia, and the PAT signal was recorded for another 5 min. The contralateral finger was used to control for systemic changes. Two variables from PAT were used: the first is mean basal pulse amplitude (BPA) that reflects the basal vascular tone and is calculated by logarithmically transforming the mean values of the basal pulse amplitudes of both arms, and for which a higher value reflects worse endothelial function. The second is PAT ratio, which reflects the response to reactive hyperemia for which a lower value reflects a more impaired response. The reproducibility of PAT in the ELSA-Brasil study was previously reported.^20^

In this study, BPA and PAT ratio were categorized into sex-specific quartiles, as women were observed to have better endothelial function, characterized by lower BPA and higher PAT ratio **(Supplementary Figure 1)**. We used the first quartile as the reference category for BPA and the fourth quartile as the reference category for PAT ratio, for both sexes.

### Outcome Variable: Type 2 Diabetes Mellitus

The response variable was the time until the first diagnosis of diabetes occurring after study entry up to December 31^st^, 2019. Participants who died of unrelated causes or who were lost to follow-up before an event was observed were censored at the time of last observation. Diabetes was dichotomously defined in participants who: (i) reported a medical diagnosis of diabetes or current use of medication for diabetes, or (ii) had a laboratory measurement reaching the thresholds for fasting plasma glucose (FPG) (≥ 7.0 mmol/L; 126 mg/ dL), 2 h post-load glucose (PG) (≥ 11.1 mmol/L; 200 mg/dL), or HbA1c (≥ 48 mmol/mol; 6.5%).^21,22^ We excluded prevalent diabetes at baseline and ascertained incident diabetes at follow-up visits based on the same criteria. We additionally included incident cases of diabetes if the participant reported a diagnosis of diabetes on at least two annual telephone interviews after the last clinical visit.^23–25^

### Covariables

The educational level was categorized as incomplete elementary school, complete elementary school, complete secondary school, or university degree. Physical activity was assessed by the long version of the International Physical Activity Questionnaire and participants were classified as sedentary (<600) or active (>600 metabolic equivalent-min/week).^26^ Smoking status was self-reported and stratified as smoker or not smoker. The participant was classified by excessive alcohol consumption when a consumption ≥ 210g alcohol/week for men and ≥ 140g alcohol/week for women was reported. Body mass index (BMI) was calculated as weight in kilograms divided by height in square meters. Waist circumference was not included in the same models as BMI due to high collinearity, as confirmed by the *estat vce* command from Stata, which identified a correlation of 0.92 between the two variables. However, as a sensitivity analysis, we replaced BMI by WC in the fully adjusted models and in the evaluation of multiplicative interaction.

### Statistical Analysis

Characteristics of the study population at baseline visit were described as proportions for categorical variables, and as means and standard deviations or medians and interquartile range for continuous variables.

We calculated incidence rate according to sex-specific quartiles of BPA and PAT ratio. The association of the sex-specific quartiles of endothelial function variables (PAT ratio and BPA) with the time until the first diagnosis of diabetes was evaluated using Cox proportional hazard models. We demonstrated this association adjusted for sex, and age (Model 1), and then further adjusted for education (Model 2), physical activity, smoking and alcohol consumption (Model 3), and finally by BMI (or WC, as a sensitivity analysis) (Model 4). Proportional hazards assumptions were evaluated using Schoenfeld residuals and violations of proportionality were not identified.

To investigate whether the effect of the BPA/PAT ratio varied according to obesity, we included multiplicative interaction terms in the final models, considering BMI or WC^27^ both as a continuous and a dichotomous variable. However, no evidence of interaction was observed in either approach (p > 0.05). We also conducted a sensitivity analysis excluding participants with prevalent CVD.

Analysis was performed using Stata software version 14.00 (Stata Corporation, College Station, United States). Data analytic methods and study materials will be made available to other researchers for reproducing the results or replicating the procedure, from the corresponding author upon reasonable request.

## RESULTS

### Participants and descriptive data

Of the 1,535 participants with valid PAT at baseline, we excluded 214 with prevalent diabetes, 47 participants were lost to follow-up, and 6 died, resulting in a final sample of 1,268 participants (**Figure 1**). Mean age was 51±8 years, with 45% women. A high proportion of sedentary (73%) and highly educated participants were observed, the latter reflecting the work-based origin of the sample. Mean BMI was 26.4 ± 4.4 kg/m^2^ **(Table 1)**. There were no statistically significant differences in baseline characteristics according to BPA **(Table 1)** or PAT ratio (**Supplementary Table 1)**, except for BMI, which was higher in individuals with more impaired endothelial function (p<0.001).

**Figure 1.**
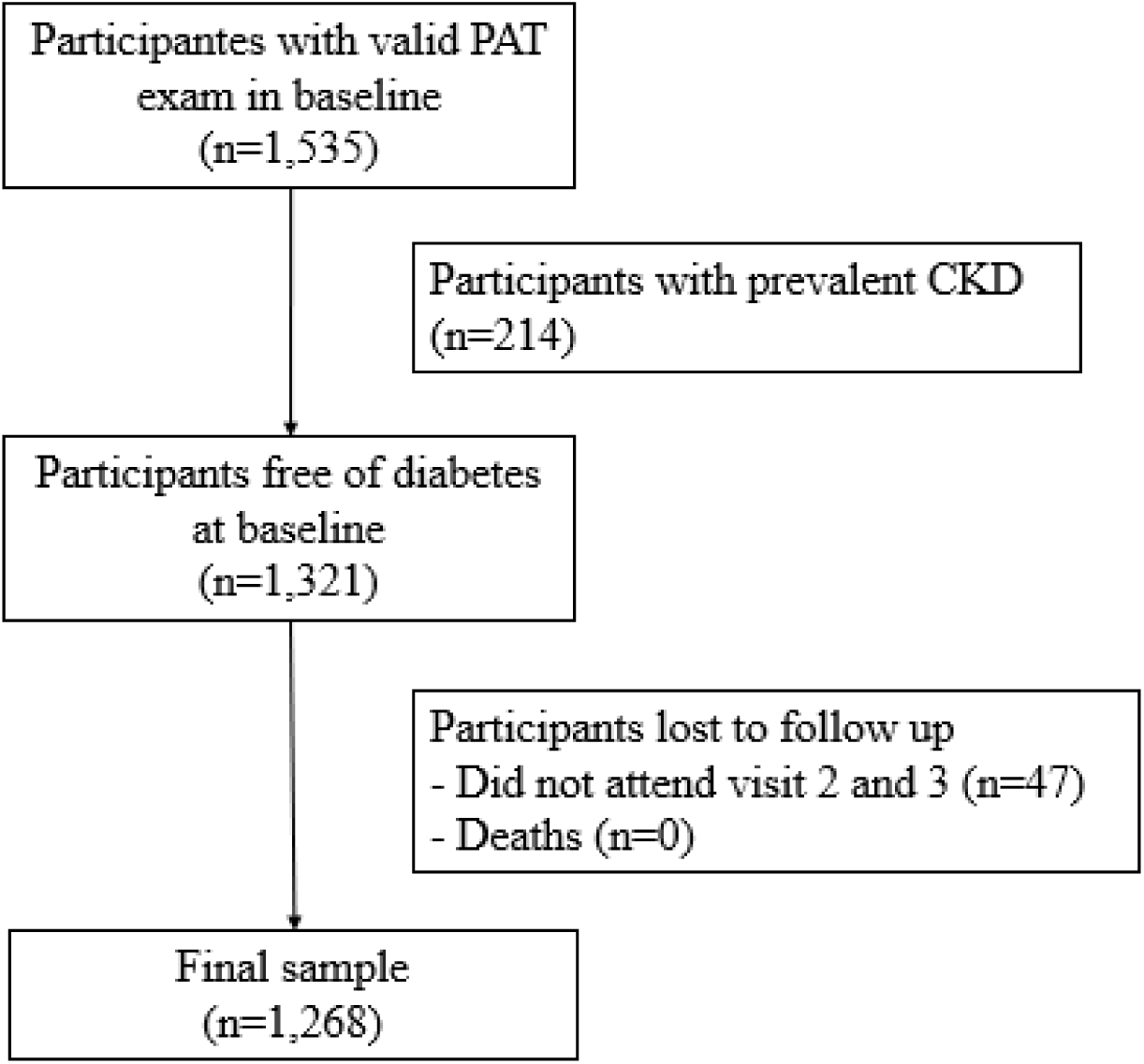
Flowchart of the study sample in present analysis. (N = 1268).

**Table 1.**
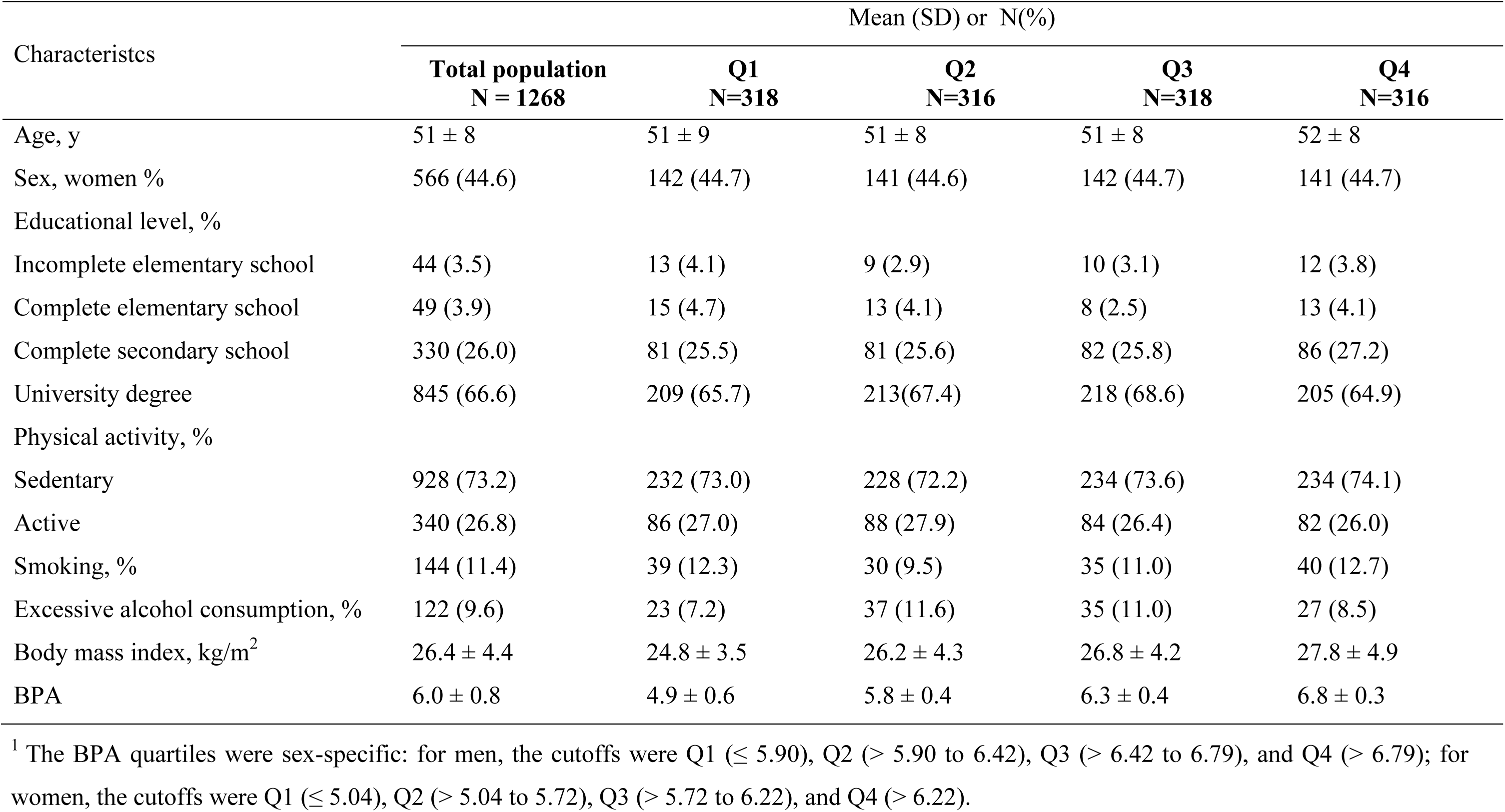
Characteristics of study participants at baseline (2008–2010), according to sex-specific quartiles^1^ of mean basal pulse amplitude (BPA). The Brazilian Longitudinal Study of Adult Health (ELSA-Brasil), N =1,268.

| Characteristics | Mean (SD) or N(%) |  |  |  |  |
| --- | --- | --- | --- | --- | --- |
|  | Total population<br>N = 1268 | Q1<br>N=318 | Q2<br>N=316 | Q3<br>N=318 | Q4<br>N=316 |
| Age, y | 51 ± 8 | 51 ± 9 | 51 ± 8 | 51 ± 8 | 52 ± 8 |
| Sex, women % | 566 (44.6) | 142 (44.7) | 141 (44.6) | 142 (44.7) | 141 (44.7) |
| Educational level, % |  |  |  |  |  |
| Incomplete elementary school | 44 (3.5) | 13 (4.1) | 9 (2.9) | 10 (3.1) | 12 (3.8) |
| Complete elementary school | 49 (3.9) | 15 (4.7) | 13 (4.1) | 8 (2.5) | 13 (4.1) |
| Complete secondary school | 330 (26.0) | 81 (25.5) | 81 (25.6) | 82 (25.8) | 86 (27.2) |
| University degree | 845 (66.6) | 209 (65.7) | 213(67.4) | 218 (68.6) | 205 (64.9) |
| Physical activity, % |  |  |  |  |  |
| Sedentary | 928 (73.2) | 232 (73.0) | 228 (72.2) | 234 (73.6) | 234 (74.1) |
| Active | 340 (26.8) | 86 (27.0) | 88 (27.9) | 84 (26.4) | 82 (26.0) |
| Smoking, % | 144 (11.4) | 39 (12.3) | 30 (9.5) | 35 (11.0) | 40 (12.7) |
| Excessive alcohol consumption, % | 122 (9.6) | 23 (7.2) | 37 (11.6) | 35 (11.0) | 27 (8.5) |
| Body mass index, kg/m<sup>2</sup> | 26.4 ± 4.4 | 24.8 ± 3.5 | 26.2 ± 4.3 | 26.8 ± 4.2 | 27.8 ± 4.9 |
| BPA | 6.0 ± 0.8 | 4.9 ± 0.6 | 5.8 ± 0.4 | 6.3 ± 0.4 | 6.8 ± 0.3 |

<sup>1</sup> The BPA quartiles were sex-specific: for men, the cutoffs were Q1 ( $\leq 5.90$ ), Q2 ( $> 5.90$ to $6.42$ ), Q3 ( $> 6.42$ to $6.79$ ), and Q4 ( $> 6.79$ ); for women, the cutoffs were Q1 ( $\leq 5.04$ ), Q2 ( $> 5.04$ to $5.72$ ), Q3 ( $> 5.72$ to $6.22$ ), and Q4 ( $> 6.22$ ).

### Outcome data

The mean follow-up time was 8.8 years (SD=2.1), ranging from 0.2 to 10.2 years. We identified 159 new cases of diabetes. The incidence rates for diabetes varied from 6.5 (95%CI 4.0-10.2) in the first quartile to 19.7 (95%CI 15.1-25.8) in the fourth quartile for BPA and from 7.2 (CI 95% 4.7-11.1) in the fourth quartile to 15.3 (CI 95% 11.4-20.6) in the first quartile of PAT ratio (**Table 2)**.

**Table 2.**
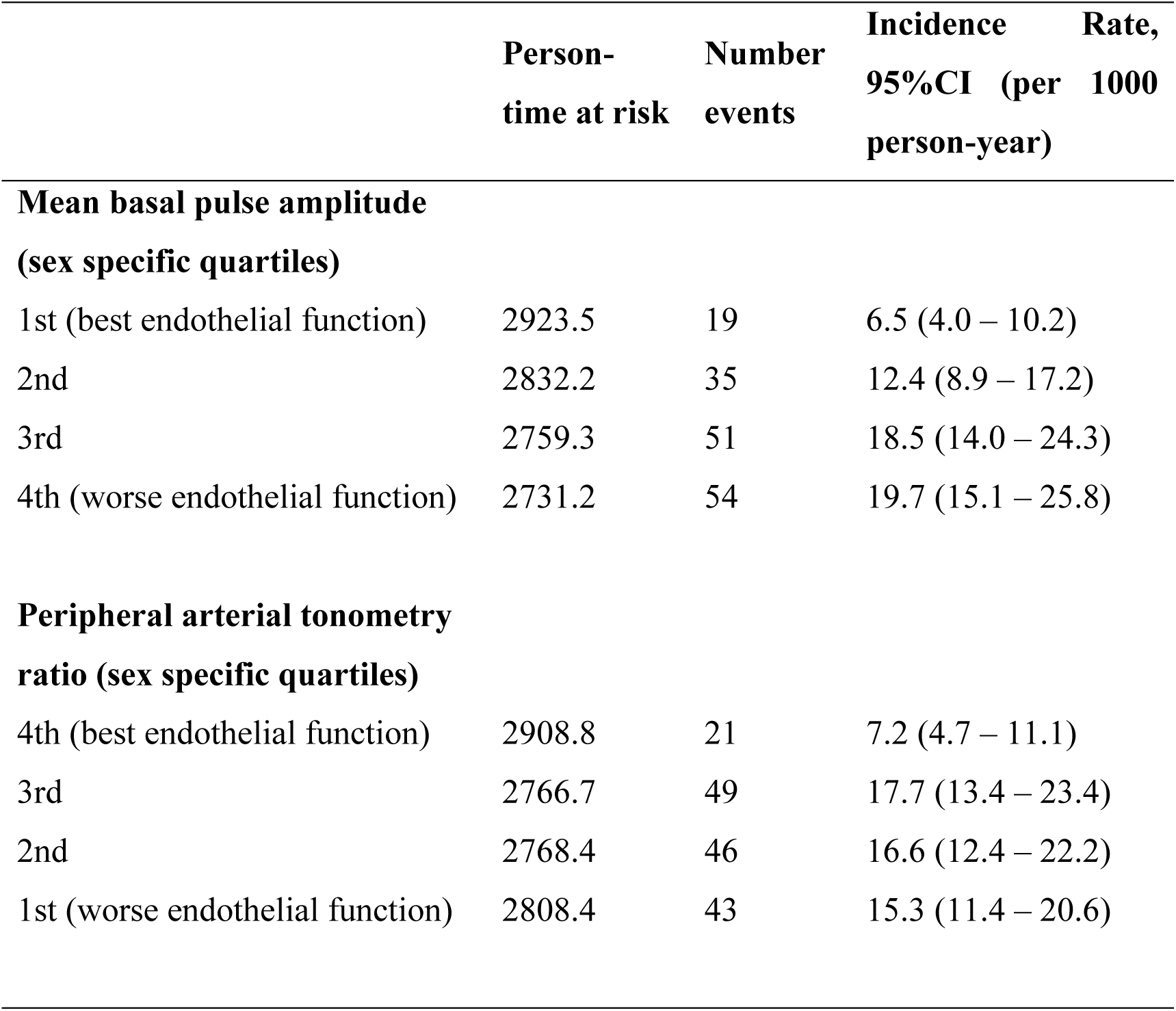
Incidence rate of type II diabetes according to sex-specific quartiles of mean basal pulse amplitude and peripheral arterial tonometry (PAT) ratio. The Brazilian Longitudinal Study of Adult Health (ELSA-Brasil, 2008-2019)

In the minimally adjusted model (Model 1), we found that the incidence of diabetes increased as BPA levels increased, showing a clear dose-response gradient. However, in the final model, after adjusting for BMI or WC, no dose-response gradient was observed **(Table 3)**. Nevertheless, the association remained statistically significant. Compared to the first quartile of BPA (indicating the best endothelial function), individuals in the second quartile were 1.68 times (95% CI 0.96 – 2.95, p 0.07) more likely to experience incident diabetes. This increased in subsequent quartiles, with 2.42 times (95% CI 1.42 – 4.12, p<0.05) higher likelihood in third quartile and 2.13 times (95% CI 1.24 – 3.66, p<0.05) higher in the fourth quartile **(Table 3)**. Similar results were found in the final model for PAT ratio. Compared to the fourth quartile, the risk of developing diabetes increased in 2.20 (95% CI 1.32 – 3.68, p< 0.05) in third quartile. In the second and first quartiles was observed a risk of 2.04 (95% CI 1.22 – 3.42, p<0.05) and 1.54 (95% CI 0.90 – 2.60, p 0.11), respectively. The variables were also evaluated together in the same model, with no changes in the results. When BMI was replaced by WC in the fully adjusted models, the results were similar (**Supplementary Table 3**). **Figure 2** shows the unadjusted cumulative incidence of type II diabetes according to BPA and PAT ratio sex-specific quartiles.

**Figure 2.**
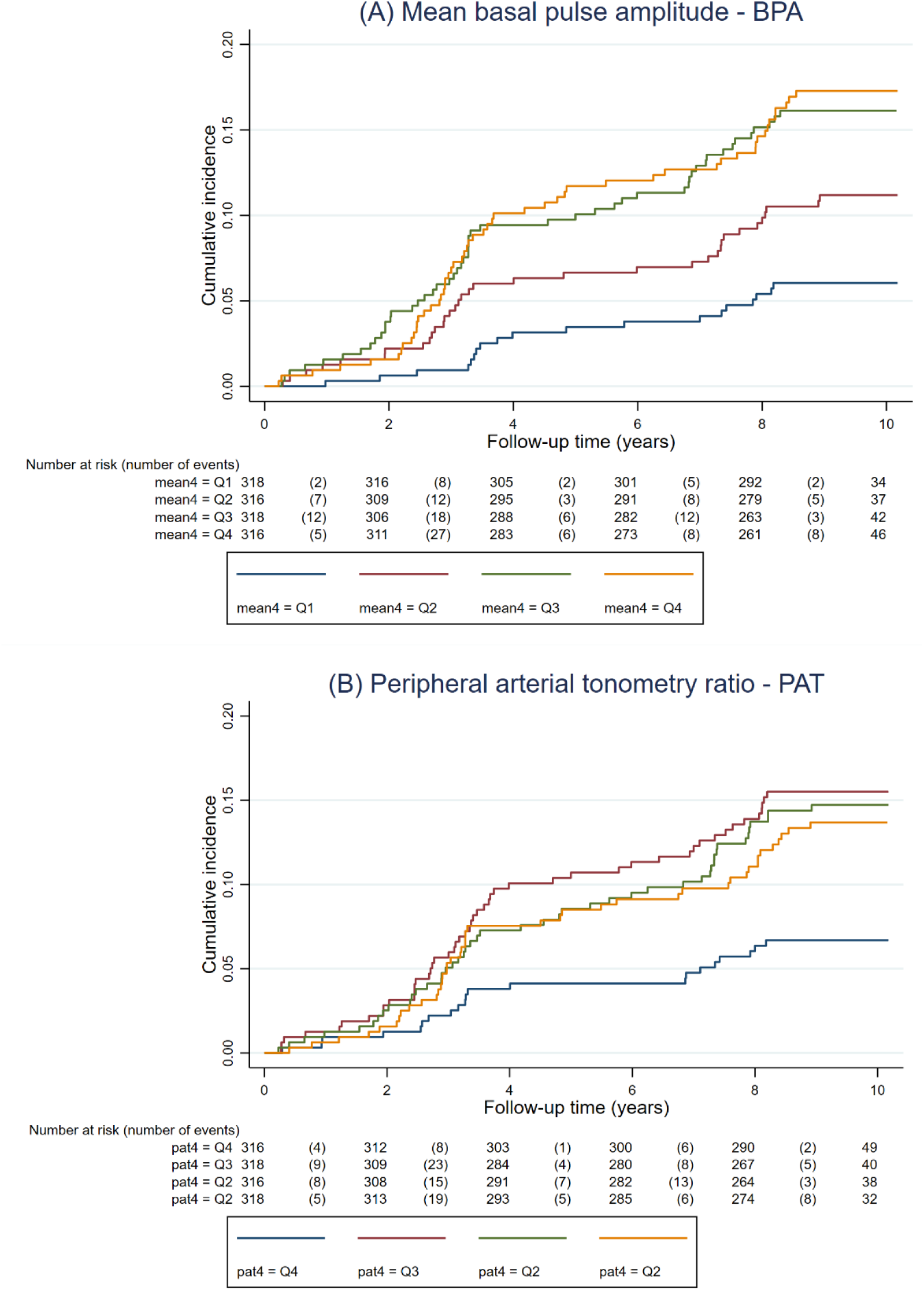
Cumulative incidence of type II diabetes according to mean basal pulse amplitude (A) and peripheral arterial tonometry ratio (B) sex-specific quartiles. The Brazilian Longitudinal Study of Adult Health (ELSA-Brasil, 2008-2019). N=1,268.

**Table 3.** Adjusted cox regression results for mean basal pulse amplitude (BPA) and peripheral arterial tonometry (PAT) ratio in sex-specific quartiles. The Brazilian Longitudinal Study of Adult Health (ELSA-Brasil, 2008-2019).

|  | **Model 1** | **Model 2** | **Model 3** | **Model 4** |
| --- | --- | --- | --- | --- |
|  | **HR (95%CI)** | **HR (95%CI)** | **HR (95%CI)** | **HR (95%CI)** |
| **Mean basal pulse amplitude** |  |  |  |  |
| **(sex specific quartiles)** |  |  |  |  |
| 1st (better endothelial function) | Ref. | Ref. | Ref. | Ref. |
| 2nd | 1.90 (1.09 – 3.33)* | 2.29 (1.10 – 3.45)* | 1.99 (1.14 – 3.49)* | 1.68 (0.96 – 2.95) |
| 3rd | 2.79 (1.65– 4.73)** | 3.86 (1.67– 4.78)** | 2.88 (1.70 – 4.88)** | 2.42(1.42– 4.12)* |
| 4th (worse endothelial function) | 2.93 (1.74 – 4.95)** | 4.05 (1.75 – 4.98)** | 2.98 (1.76 – 5.02)** | 2.13 (1.24 – 3.66)* |
| **Peripheral arterial tonometry** |  |  |  |  |
| **ratio (sex specific quartiles)** |  |  |  |  |
| 4th (better endothelial function) | Ref. | Ref. | Ref. | Ref. |
| 3rd | 2.44 (1.46 – 4.07)** | 2.45 (1.47 – 4.09)** | 2.46 (1.45 – 4.10)** | 2.21 (1.30 – 3.68)* |
| 2nd | 2.27 (1.35 – 3.80)* | 2.29 (1.36 – 3.83)* | 2.28 (1.36 – 3.81)* | 2.04 (1.22– 3.42)* |
| 1st (worse endothelial function) | 2.06 (1.22 – 3.47)* | 2.08 (1.23 – 3.50)* | 2.04 (1.21 – 3.45)* | 1.54 (0.90 – 2.61) |

Model 1: sex and age
Model 2: Model 1+ education
Model 3: Model 2+ physical activity, smoking, and excessive alcohol consumption
Model 4: Model 3+ body mass index
\*p<0.05 \*\* p≤ 0.001

We found no evidence of multiplicative interaction between BMI or WC and endothelial function variables in their association with incident diabetes. Sensitivity analysis excluding participants with prevalent CVD was performed, and the results were similar as showed in **Supplementary Table 2**.

## DISCUSSION

In the present study, we evaluated the longitudinal association between endothelial function and incident diabetes in a large cohort of Brazilian adults. Prior studies have described the association of digital vascular function with incident diabetes in individuals of European descent,^6,7,28^ who have different clinical and epidemiological characteristics compared to the Brazilian population.^8^ We herein demonstrate similar associations of more impaired endothelial function with incident diabetes after 8.8 years of follow-up in individuals from Brazil, a middle-income country, where demographic and nutritional shifts are increasing the prevalence of metabolic risk factors, particularly diabetes and obesity, which threaten to rise the incidence of CVD.^3,4^ We also observed that there was no effect modification by BMI category or WC in the association of endothelial function variables and incident diabetes, suggesting that that endothelial function is associated with diabetes independently of obesity.

After adjustments, we found that the risk of diabetes increased significantly in those with more impaired endothelial function, and individuals in the fourth quartile of BPA had a 105% higher risk of diabetes compared to those in the first quartile of BPA. Our findings are consistent with previous results from the Gutenberg Heart Study (GHS), a cohort study conducted in Europeans,^14^ which demonstrated the reactive hyperemia index (RHI) was independently associated with an increased risk of type 2 diabetes by 16% (95% CI, 1.01–1.34; P=0.041) per 1 SD. Furthermore, BPA was independently predictive of future development of diabetes (RR, 1.17; 95% CI, 1.02–1.33; P=0.022). These results corroborate what we observed in direction and significance, even though there were methodological differences in our studies, precluding direct comparison of effect sizes: we used PAT ratio instead of RHI; we used different distribution of data and statistical model, as the associations of our explanatory variables with diabetes were non-linear; we adjusted for a smaller number of covariables that were considered confounding factors in our conceptual model.

Some studies have also evaluated whether endothelial dysfunction can predict diabetes in different scenarios.^29–34^ To compile these results, in 2012, Muris *et al* published a meta-analysis of 13 studies demonstrating that various estimates of microvascular dysfunction were associated with incident diabetes, including plasma samples of E-selectin, vascular cell adhesion molecule-1 (VCAM-1), intercellular adhesion molecule-1(ICAM-1), and Von Willebrand factor (vWF) analyzed by use of ELISAs; peripheral vascular reactivity assessed by a dose-response curve to intra-arterial infusion of acetylcholine; retinal diameters evaluated by the use of fundus photos and microalbuminuria. However, only one of these studies evaluated vascular function, and the method used was invasive, and in large conduction vessels.^12^ Our study brings novelty by investigating a non-invasive method in the community-based and racially diverse ELSA-Brasil study.

We also observed that BPA was more consistently associated with incident diabetes than PAT ratio. This finding has also been previously reported with other cardiovascular risk factors,^6–8^suggesting that these risk factors may be more associated with structural changes than physiological responses, reflected by the higher basal vascular tone, expressed by BPA. A question already raised was that, given the strong correlation between the PAT ratio and BPA, part of the PAT ratio values could be a result of an impaired BPA and not a response to hyperemia. The influence of the resting pulse amplitude may be a physiological effect given that the already higher pulse amplitude could preclude vasodilation, or a mathematical calculation effect, because PAT ratio is calculated using BPA in the denominator.^5^

We did not confirm our hypothesis that overweight/obesity would amplify the effect of endothelial dysfunction on diabetes risk, as we found that BMI and WC did not modify the relationship between endothelial dysfunction and diabetes risk. However, we observed that after adjusting for those, the association between BPA and diabetes risk diminished, suggesting that part of the effect of endothelial dysfunction on diabetes risk is explained by obesity. This could be explained by two mechanisms: 1) Obesity may act as a common cause of both endothelial dysfunction and diabetes, thereby confounding the observed relationship between endothelial function and diabetes; 2) endothelial dysfunction could increase the risk of obesity, which in turn could contribute to the development of diabetes, with BMI and waist circumference potentially serving as a mediators in the endothelial dysfunction diabetes association. In fact, longitudinal studies have shown that obesity is an important predictor of endothelial dysfunction,^35, 36^ with support the first hypothesis. On the other hand, previous studies hypothesize that alterations in endothelial function could be sufficient to accelerate obesity.^37,38^ Some molecules are considered key and capable of modulating systemic metabolism, including transcription factors (P53, PGC1a, FOXOs and NF-kB), pro-angiogenic signals (VEGD, VEGFR2, ANG2/TI2, DLL4/NOTCH1), components of the insulin cascade (IRS-2, insulin receptor) and mediators of fatty acid transport (NOTCH1, VEGF-B/NRP1 and CD36) ^39–46^

One of the insulin’s actions involves the production of the vasodilator NO, increasing blood flow and glucose uptake by skeletal muscle. At the cellular level, the balance between phosphatidylinositol 3-kinase, representative of the insulin pathway that regulates endothelial production of NO (PI3K-NO), and mitogen activation of protein kinase (MAPK), representative of the pathway that regulates the secretion of the vasoconstrictor endothelin-1 (MAPK-ET-1), determines the vascular response to insulin. Endothelial insulin resistance is typically accompanied by a reduced PI3K-NO pathway and an intact or increased MAPK-ET-1 pathway contributing to endothelial dysfunction.^47^ However, the interaction between glucose metabolism and endothelial dysfunction is more complex and involves several feedback mechanisms. In the arterial microcirculation, endothelial dysfunction can impair the insulin’s function and redirect blood flow from non-nutritive to nutritive capillaries, reducing muscle glucose uptake.^48–50^ Pancreatic microvascular dysfunction can cause apoptosis of βcells in the pancreas, reducing insulin secretion from the pancreas and thus leading to hyperglycemia^51^ which has been associated with endothelial dysfunction.^52^ Furthermore, microvascular dysfunction may act as an intermediate step linking obesity, low physical activity, and chronic low-grade inflammation to insulin resistance,^53^ although the effect modification by obesity status was not observed in this analysis.

Our study has limitations. Because the endothelial function evaluation was a substudy of ELSA-MG, our sample size was smaller than that of the ELSA-Brasil study, precluding stratified analysis for incident pre-diabetes and diabetes. Since ELSA-Brasil is an observational study, we cannot rule out the possibility of residual confounding by the variables included in the adjustment model, nor can we completely exclude the possibility of omitted confounding variables. Regarding PAT exam, it evaluates microvascular endothelial function, which has the advantage of being noninvasive and operator independent, but may not be used interchangeably with other measures of vascular function.^8,9^ Another issue is that BPA and PAT ratio do not have cutoffs that could be used to categorize the endothelial function variables. To overcome this issue, we studied the variables continuously and in sex-specifiquartiles before deciding how to analyze them. We recognize that the absence of a dose-response relationship between AMB/PAT ratio and diabetes are also a limitation to the interpretation of results. Lastly, we acknowledge that BMI alone is not the ideal measure of excess adipose tissue,^27^ and that WC may be more related to metabolic risk^54,55^ We chose to use BMI as it is more frequently used in clinical practice. To overcome this limitation, we performed a sensitivity analysis using WC instead of BMI and the results were largely unchanged. Strengths of the present study include the use of a robust statistical model demonstrating the association between endothelial dysfunction and incident diabetes in a diverse population with a high prevalence and incidence of diabetes, in which this association had not yet been demonstrated, and the comprehensive and rigorous assessment of covariables and ascertainment of outcomes.

## CONCLUSION

Our findings support the hypothesis that impaired endothelial function in the microvasculature occurs before the onset of type 2 diabetes, corroborating data observed in the European population, and advancing the field by showing that this association is not modified by obesity status. Our findings may reflect that endothelial dysfunction could be part of a cyclical mechanism that will feedback metabolic endocrine changes, predisposing to diseases such as diabetes and, subsequently, CVDs. These findings may have implications for the understanding of cardiometabolic diseases in the context of pathophysiology, prognosis and treatment. ELSA-Brasil has the possibility to continue exploring the topic to help elucidate these relationships.

## Data Availability

All data referred to in the manuscript are available upon request from the corresponding author.

